# Detection of transmission change points during unlock-3 and unlock-4 measures controlling COVID-19 in India

**DOI:** 10.1101/2020.11.17.20233221

**Authors:** Manisha Mandal, Shyamapada Mandal

## Abstract

Documentation in scientific literature is not available on prospective evaluation of the efficiency of the unlock measure related to COVID-19 transmission change points in India, projecting the infected population, planning suitable measures related to future interventions and lifting of restrictions so that the economic settings are not damaged beyond repair. We have applied SIR model and Bayesian approach combined with Monte Carlo Markov algorithms on the Indian COVID-19 daily new infected cases from 1 August 2020 to 30 September 2020. We showed that the COVID-19 epidemic declined after implementing unlock-4 measure and the identified change-points were consistent with the timelines of announced unlock-3 and unlock-4 measure, on 1 August 2020 and 1 September 2020, respectively, effectiveness of which were quantified as the change in both effective transmission rates (100% reduction) and the basic reproduction number attaining 1, implying measures taken to control and mitigate the COVID-19 epidemic in India managed to flatten and recede the epidemic curve.

## 1. Introduction

To contain COVID-19 spread in India, strong phasic lockdowns were implemented leading to reduction of human contact to a maximum 55%, and 34% at the end of lockdown, followed by stratified unlock measures with gradual return to activities, controlling social contacts to 19% reduction, as on 30 September, 2020 (IHME, 2020). India, currently is the world’s second-worst-hit country with nearly 11.7 million COVID-19 infections including more than 98,000 deaths, as on 30 September 2020 (COVID19India, 2020). Until COVID-19 is completely eradicated, and effective treatment or vaccine become available, non-pharmaceutical intervention policies are the key public health options to control the epidemics (Varghese et al., 2020). During the evolution of COVID-19, India implemented lockdowns in four phases from 24 March to 31 May 2020 as containment and mitigation measure, followed by unlocks in four phases from 1 June to 30 September 2020, featured by conditional relaxations of restrictions outside containment zones in graded manner, to minimise the negative economic and social consequences of strict lockdown measures (MOHWF, GoI, 2020). With the escalating case numbers and prolonged COVID-19 epidemic situation in India, the present study is an investigation to determine, if within the currently implemented non-pharmaceutical strategies, taking into account the simulated stochastic SIR model of transmission dynamics, is effective in curbing the spread of COVID-19. For this purpose, we made posterior inference on transmission rate *λ*, recovery rate *μ*, reproduction number *R*_0_, number of initially infected people *I*_0_, reporting delay *D*, width of liklihood *σ* between observed daily infected cases and its best fit estimates, effective transmission rate *λ*^*\**^ = λ − μ, based on data-driven likelihood updates of prior settings. We determined also the change-points in disease transmission and investigated the effectiveness of unlock-3 and unlock-4 measures, with respect to their strength, timing and duration. We used an open source probabilistic programming in Python code PyMC3 with theano to compute gradients via automatic differentiation variational inference (ADVI), and followed model interpretation on German COVID-19 data (Dehning et al., 2020), based on GitHub repository (Priesemann-Group, 2020), to analyse the recent COVID-19 pandemic situation in India with emphasis on unlock-3 and unlock-4 measure.

## 2. Methods

### 2.1. Data sources

The data on ongoing new daily and cumulative COVID-19 cases in India were retrieved from Johns Hopkins University Centre for Systems Science and Engineering dashboard, up to September 30, 2020 (CSSE, 2020). The codes for this research article pertaining to the analysis of unlock-3 and unlock-4 situation in India was run on Jupyter notebook using PymC3=3.8 and was based on GitHub repository (Priesemann-Group, 2020), by importing python-based data analysis toolkit (pandas); libraries for working with arrays (numpy), plotting (matplotlib), scientific and technical computing (scipy), and multi-dimensional arrays (theano); modules including Basic date and time types (datetime), System-specific parameters and functions (sys), and Python object serialization (pickle); package for Bayesian statistical modeling and probabilistic machine learning with MCMC and ADVI algorithms (PymC3) (Kucukelbir et al., 2007).

### 2.2. SIR model

The SIR model was based on time-varying cumulative number of COVID-19 cases, where the total population size (*N*) was categorized into three mutually exclusive infection levels, assuming that any infectious person (*I*), is likely to contact any susceptible person (*S*), and later recovered (*R*), so that *N* = *S* + *I* + *R*. The dynamics of the pandemic in India was modelled using the following three differential equations:

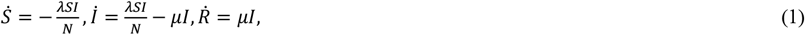

where *λ* represents the transmission rate of the infected people to infect susceptible people and *μ* denotes the recovery rate of the infected people to recover (Kermack et al., 1927). This is solved by using a forward finite-difference scheme (Carcione et al., 2014):

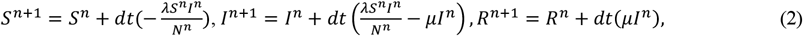

where *n* is a natural number which divides time *t* in *n* discrete *dt* time steps, *t* = *ndt*.

The fraction of maximum number of infected people, 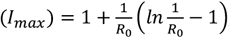, where 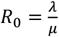 and the fraction of people remaining susceptible to infection (*S*_*inf*_) is related to *R*_0_ by: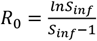. The overall infection attack rate (*IAR*) defined as the fraction of the population that eventually becomes infected is related to *R*_0_ by: 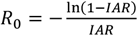

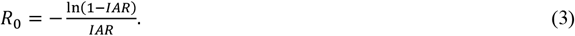

### 2.3. SEIR model

The SEIR model is an extension of the SIR with an added exposure (*E*) period due to the reported incubation period of COVID-19 during which individuals are not yet infectious (Hethcote, 2000). The SEIR models the total population size (*N*) divided into four mutually exclusive infection stages, *N* = *S* + *E* + *I* + *R*, and is based on following differential equations:

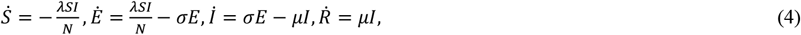

where, σ is the rate at which individuals in incubation become infectious. The differential equation is solved as (Carcione et al., 2014):

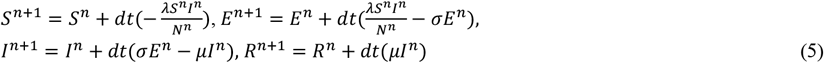

### 2.4. Model inclusions

A reporting delay D, was incorporated in becoming infected (*I*^*new*^) and being reported, such that the daily reported cases *R*_*t*_ at any time t was given by (Dehning et al., 2020), 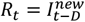. To examine if there was any weekend effect on daily reported case numbers, a periodic sine function was assigned to the reporting fraction *f*(*t*) expressed as,

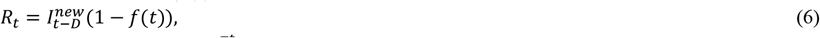

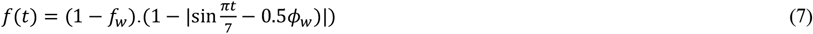

where *f*_*w*_ and *ϕ*_*w*_ are the weekly modulation amplitude and phase respectively (Dehning et al., 2020).

### 2.5. Bayesian Inference

For a statistical model, *p*(*x*|*θ*), that reflects our beliefs about *x* given*θ*, with the prior distribution *π*(*θ*),on an observed data*D*_*n*_ = {*X*_1_, …, *X*_*n*_}, the posterior distribution is expressed as (Bayes, 1763; Box et al., 1992):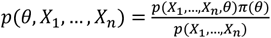

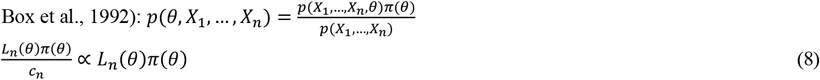

where, 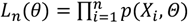 is the likelihood function.

The likelihood is a measure of the goodness of fit between model prediction and the observed data on reported case numbers, applied hereby using Student-t distribution. The evidence

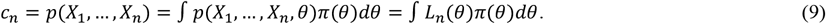

The Bayesian posterior interval estimate for *a* and *b, α ∈* (0,1), *C* = (*a, b*), is given by,

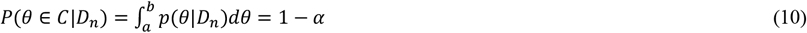

The Bayesian predictive distribution is

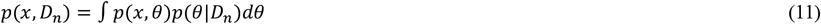

The inferences about a function *τ* = *g*(*θ*), so that cumulative distribution function for *τ*is *H*(*t,D*_*n*_) =*P*(*g*(*θ* ≤*t,D*_*n*_)

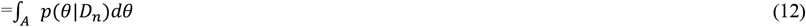

where *A* = (*θ*: g(θ) ≤ t); the posterior density is *p*(*τ, D*_*n*_) = *H*^’^(*τ, D*_*n*_).

### 2.6. Priors

The prior distribution settings for the model parameter estimation were made by incorporating LogNormal values of λ, μ, and D and half-Cauchy distribution for I_0_, and σ (Table 1 and 2). The priors on change points in transmission rate were based on announcements of applied intervention including unlock-3 on 1 August 2020 and unlock-4 on 1 September 2020.

**Table 1.**
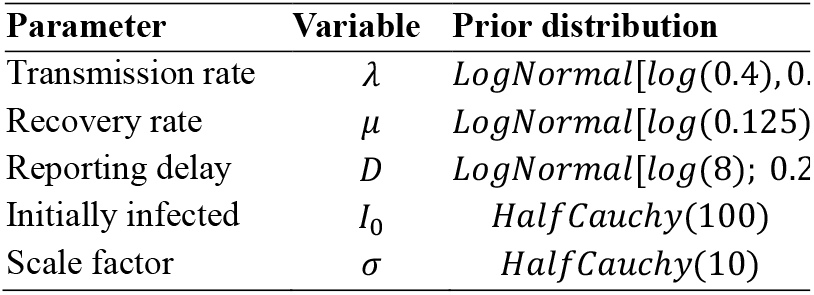
Prior distribution settings for India unlock-3 and unlock-4 SIR model parameters with fixed transmission rate.

**Table 2.** Prior distribution settings for SIR model parameters with changing transmission rates and weekend reporting factor.

| Parameter | Variable | Prior distribution |
| --- | --- | --- |
| Change points | $t_1$ | 01 August 2020 |
| | $t_2$ | 01 September 2020 |
| Change duration | $\Delta t_i$ | $LogNormal[\log(3), 0.3]$ |
| Spreading rate | $\lambda_0$ | $LogNormal[\log(0.4), 0.5]$ |
| | $\lambda_1$ | $LogNormal[\log(0.16), 0.5]$ |
| | $\lambda_2$ | $LogNormal[\log(0.15), 0.5]$ |
| Recovery rate | $\mu$ | $LogNormal[\log(0.125), 0.2]$ |
| Reporting delay | $D$ | $LogNormal[\log(8), 0.2]$ |
| Initially infected | $I_0$ | $HalfCauchy(100)$ |
| Scale factor | $\sigma$ | $HalfCauchy(10)$ |
| Weekly modulation amplitude | $f_w$ | $\beta(\text{mean} = 0.7, \text{standard deviation} = 0.17)$ |
| Weekly modulation phase | $\phi_w$ | $VonMises(\text{mean} = 0, k = 0.01)$ |

### 2.7. Markov Chain Monte Carlo (MCMC) sampling

This method is essentially a Monte Carlo sampling with multiple Markov chains, used to approximate the posterior distribution of model parameters by including ADVI, 1000 tuning steps with NUTS (No U Turn Sampling) algorithm (Hoffman et al., 2014) for each of four chains, and R-hat diagnostics for equilibrated chain convergence of model parameters (Vehtari et al., 2019). A sequence of random variables{*X*_1_, …, *X*_*n*_}, on a discrete state apace is called a Markov chain if

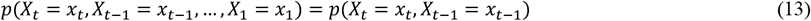

We wanted to find a setting of a parameter *x ∈ R*, such that the expectation *h*(*x*) ≡ *E*_*t*_(*H*_*t*_, *x*) = 0, the updates were applied as (Hoffman et al., 2014):

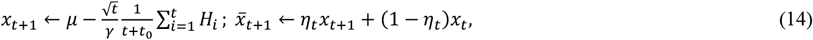

where t is iteration, *η*_*t*_is the step size schedule, *μ* is a freely chosen point that the iterates *x*_*t*_ are shrunk towards, *γ* > 0 is a free parameter that controls the amount of shrinkage towards *μ, t*_0_ ≥ 0 is a free parameter that stabilizes the initial iterations of the algorithm, *η*_*t*_ ≡ *t*^−*κ*^ is a step size schedule satisfying the conditions, ∑_*t*_ *η*_*t*_ = ∞; ∑_*t*_ *η*_*t*_^2^ < ∞. The step size parameter was set for NUTS using stochastic optimization with vanishing adaptation of the primal-dual algorithm. For each iteration we defined the statistic 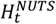 and its expectation when the chain reached equilibrium as (Hoffman et al., 2014):

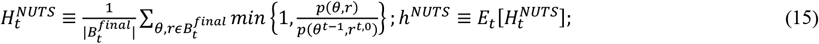

where 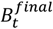 is the set of all states explored during the final doubling of iteration *t* of the Markov chain; *θ*^*t*−1^, *r*^*t*,0^ are the initial position and sampled momentum for the *t*^*th*^ iteration of the Markov chain; *H*^*NUTS*^ is the average acceptance probability. We applied in the above updates equation: *H*_*t*_ ≡ *δ* − *H*^*NUTS*^ and *x* ≡ *log ϵ* for the step size *ϵ* to combine *h*^*NUTS*^ = *δ* for any *δ ∈* (0,1).

### 2.8. Model comparison

For model fit and comparison using MCMC, following computations were made using *LOO* (leave one out) package in PyMC3: the Bayesian *LOO* estimate of the expected log pointwise predictive density (*ELPD*-*LOO*) for a new point, standard error (*SE*) of *ELPD*-*LOO*, the difference between *ELPD*-*LOO* and the non-cross-validated log posterior predictive density (*pLOO*)interpreted as the effective number of parameters (Vehtari et al., 2017). Lower *LOO* scores indicated better consistency between models. *LOO* scores with *SE* < 1 represented model compatibility while *LOO* scores *SE* > 1 indicated mismatch between the models.

For data 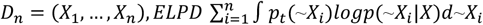, where *p*_*t*_(∼*X*_*i*_)is the distribution of the true data generating process for ∼*X*_*i*_, which is approximated by cross-validation with

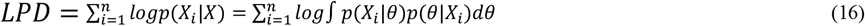

The *LPD* computed with *S* draws from a posterior distribution = 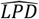 = computed log pointwise predictive density

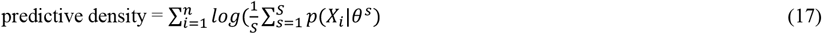

The *ELPD*-*LOO*, calculated by cross-validation by running the model *n* times is

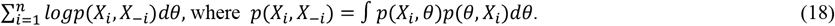

## 3. Results and Discussion

The median posterior distribution of COVID-19 epidemiological parameters using SIR model (from Eqs. 1 and 2) combined with Bayesian inference (generated by Eqs. from 8 to 15) during unlock-3 were *λ* = 0.15 (0.11 − 0.22), *μ* = 0.14 (0.10 − 0.21), *R*_0_ = 1.07 (1.05 − 1.10), *I*_0_ = 327139 (220028 − 450505), *D* = 8 days (5.5 − 11.8), *σ* = 18.7 (13.4 − 26.1), *λ*^*\**^ = 1(1 − 2), the values in bracket indicate 95% confidence-intervals *CIs* (Fig. 1), according to Eqs. from 16 to 18, *ELPD*-*LOO* = −305.07, *pLOO* = 3.04, computed from 2000 by 30 log-likelihood matrix (Table 3). The corresponding values during unlock-4 were *λ* = 0.15 (0.10 − 0.20), *μ* = 0.15 (0.10 − 0.20), *R*_0_ = 1.00, *I*_0_ = 642500 (455336 − 922684), *D* = 8.0 days (5.4 − 11.9), *σ* = 19.1 (13.5 − 26.8), *λ*^*\**^ = (−1 − 0) (Fig. 2), *ELPD*-*LOO* = −299.14, *pLOO* = 2.74, computed from 2000 by 29 log-likelihood matrix (Table 3). The median estimates of *R*_0_ decreased from 1.07 in the unlock-3 period to 1.00 in the unlock-4 period, with SIR model, indicating slowing down of the spread of the disease. The *R*_0_ was reported as 1.14 at the end of August 2020, and 1.12 in the mid of September 2020 in India, with stationary-time-series auto regressive integrated moving average model (Yadav et al., 2020). The mathematical models help to determine the effect of preventive policies against COVID-19, primarily by maintaining the reproduction number *R*_0_ < 1 to inhibit further spread of infection, whereas *R*_0_ > 1 indicate continuation of the epidemics, which fade away when the transmissibility is reduced by 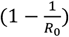 (Ferguson et al., 2020; Mandal et al., 2020; Pan et al., 2020).

**Table 3.** Bayesian LOO estimate of the ELPD for model comparison.

| Model | ELPD-LOO | p-LOO | StandardE<br>rror | n<br>log-likelihood<br>matrix |
| --- | --- | --- | --- | --- |
| SIR during unlock-3 | -305.07 | 3.04 | 4.39 | 30 |
| SIR during unlock-4 | -299.14 | 2.70 | 3.90 | 29 |
| SIR during unlock-3 and intervention evaluation | -305.39 | 3.45 | 4.58 | 30 |
| SIR during unlock-4 and intervention evaluation | -299.04 | 2.75 | 4.08 | 29 |
| SIR with 2 change points and without weekend factor | -687.67 | 6.71 | 8.00 | 68 |
| SIR with 2 change points and with weekend factor | -656.91 | 7.43 | 6.82 | 68 |
| SEIR with 2 change points and with weekend factor | -650.94 | 13.66 | 7.16 | 68 |
| SIR with 1 change point and with weekend factor | -700.05 | 6.24 | 6.14 | 68 |

**Fig. 1.**
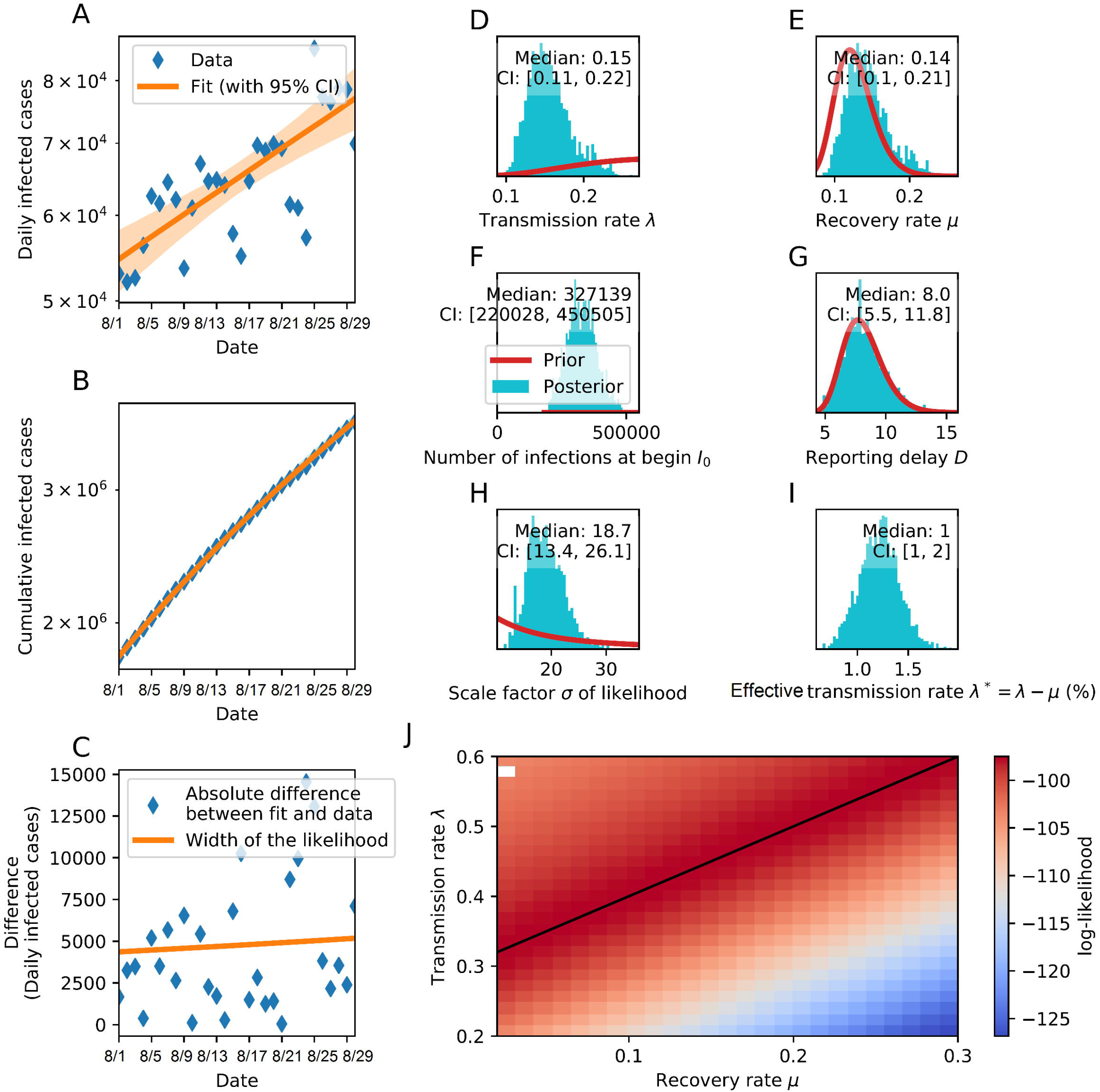
Bayesian inference of India SIR COVID-19 epidemic model parameters during unlock-3 from 1 to 31 August 2020. Exponential growth of **(A)** daily infected cases *y* = 53694.96*e*^0.01050*x*^ and **(B)** cumulative infected cases *y* = 1702723.32*e*^0.025x^with decreasing variation in rate of change of the logarithmic cumulative case, average 0.0108 (0.0104 − 0.0116); **(C)** difference in daily infected cases between fit and data; priors (red) based posterior (cyan) inference of **(D)** transmission rate *λ*, **(E)** recovery rate *μ*, **(F)** number of initially infected people *I*_0_, **(G)** reporting delay *D*, **(H)** scale factor of liklihood *σ* between observed daily infected cases (blue) and its median fit with 95% *CI* (orange), **(I)** effective transmission rate *λ*^*\**^ = *λ* − *μ*; **(J)** log-likelihood combination of transmission and recovery rates with maximum value (black line) and data non-convergence (white rectangle). Serial interval (*SI*) across India during unlock-3 was 27.43 days (26.06 − 29.25); recovery rate (*RR*) 71.9 % (76.35 − 79.16); fatality rate (*FR*) 1.93% (1.91 − 1.97); reproduction number *R*_0_1.07; *IAR* 14.2% people per million population, *I*_*max*_ = 0.00218 during unlock-3, and *S*_*inf*_ = 0.9345 after unlock-3, computed from Eqn. 3.

**Fig. 2.**
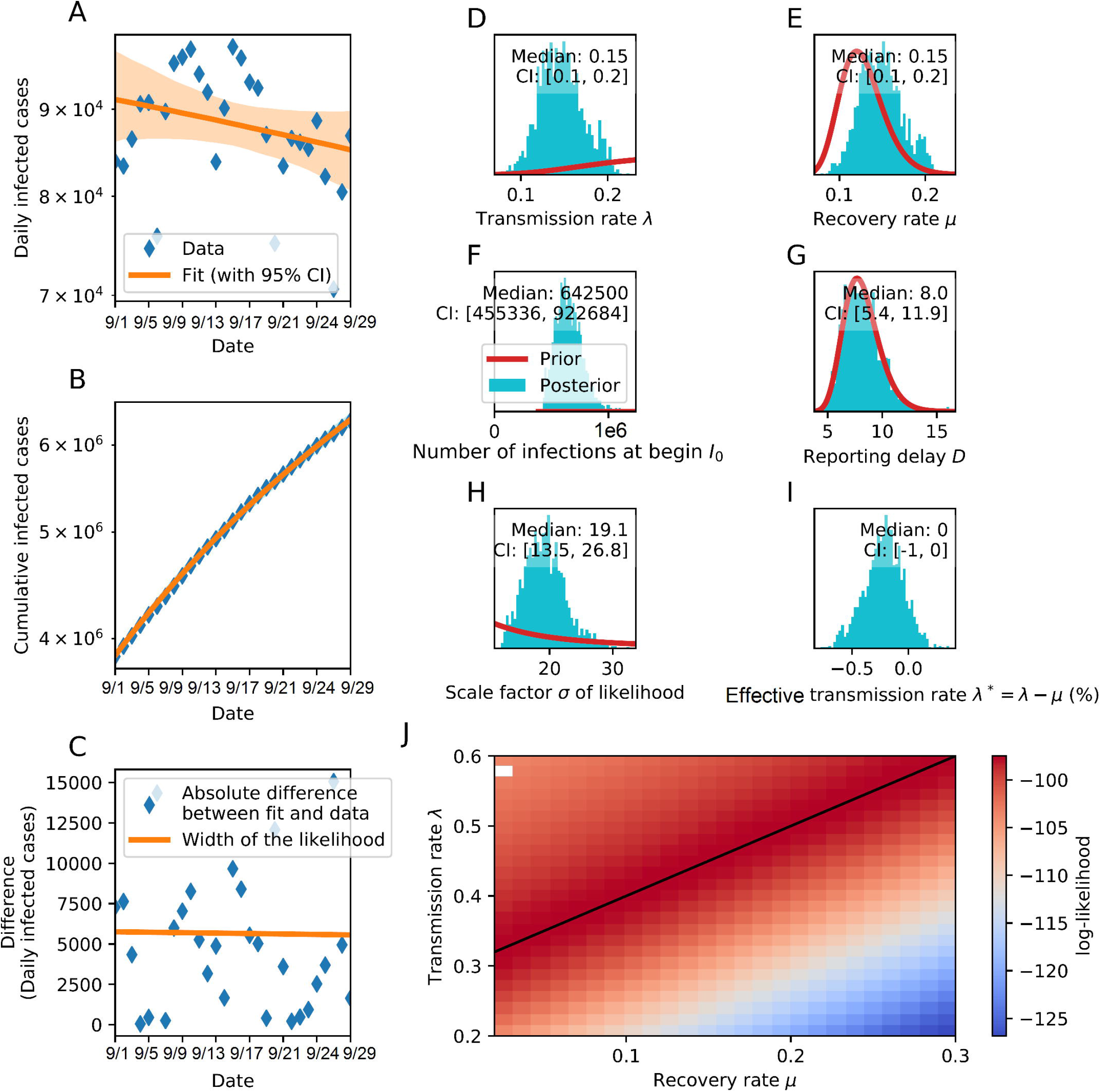
Bayesian inference of India SIR COVID-19 epidemic model parameters during unlock-4 from 1 to 30 September 2020. Implementation of unlock-4 measure was consistent with exponential **(A)** decay of daily infected cases *y* = 87133.43*e*^−0.00049x^ and **(B)** growth of cumulative infected cases *y* = 3700990.43*e*^0.018x^ with diminished rate and continued dwindling in rate of change of the logarithmic cumulative case, average 0.0080 (0.0075 − 0.0084); **(C)** difference in daily infected cases between fit and data, with declining width of liklihood; priors based posterior inference of **(D)** *λ*, **(E)** *μ*, **(F)** *I*_0_, **(G)** *D*, **(H)** *σ*, **(I)** *λ*^*\**^; (J) log-likelihood between *λ* and *μ*. Compared to unlock-3, unlock-4 strategies featured higher *SI* 37.35 days (36.43 − 42.21), enhanced *RR* 79.23 % (78.44 − 80.01), reduced *FR* 1.64% (1.62 − 1.67), decreased *R*_0_ 1.00, increased *IAR* at 18.6%, *I*_*max*_decreased to zero during unlock-4; *S*_*inf*_ increased to 1.075 after unlock-4.

The median daily COVID-19 infected cases in India at the end of unlock-3 reached ≈ 80,000, that increased about 1.09 times, to ≈ 87,500 at the end of unlock-4 (Figs. 1A and 2A), while the median cumulative infected cases increased 1.8 fold to 6.3 million at the end of unlock-4 from 3.5 million at the end of unlock-3 (Figs. 1B and 2B). The difference in daily infected cases showed approximately 1.125 fold change to 5,625 during unlock-4 on 30 September, 2020, from 5000 during unlock-3 on 31 August, 2020 (Figs. 1C and 2C). The COVID-19 daily cases increased during unlock-3 but decreased during unlock-4, though with a higher end point (Figs. 1A and 2A). The real-time daily and cumulative cases were consistent with SIR-model displaying linear-semi-logarithmic variation. A decreasing first order differences of the logarithm of the cumulative cases over time indicated exponential growth as found for India, whereas a constant trend indicated logarithmic growth of the epidemic curve as seen in the US, for the period from 17 September 2020 to 1 October 2020 (Baruah, 2020).

The priors and posteriors exhibited different *λ, I*_0_, and *σ* (Figs. 1D, 1F, 1H, 2D, 2F, and 2H) implicating informative feature of the observed data; and matched *μ* and *D*, indicating dependency of the observed data on prior informatives (Figs. 1E, 1G, 2E, and 2G), in unlock-3 and unlock-4 exhibiting exponential transmission rates. The *λ*^*\**^ became zero in unlock-4, from 10% in unlock-3 (Figs. 1I and 2I), indicating post unlock-4 effect through inhibition of new infections. The *σ*, a measure of goodness of fit between *λ* and *μ*, showed equipotential line for the maximum liklihood (Figs. 1J and 2J), implicating *λ*^*\**^ as an important and independent regulator of COVID-19 transmission dynamics.

Both daily and cumulative infected cases remain unaltered until the duration of *D* and change-point were over, beyond which both continued to rise, build on the hypothesis that ongoing unlock phase and its post-effect prevailed, but continuation of pre-unlock situation caused decline in both, the effect being more significant in the daily cases (Fig. 3A and 3D). The daily as well as cumulative case numbers showed rising trend in post unlock-3 scenario (Fig. 3A), however, the onset timings of intervention had no effect on case numbers (Fig. 3B). With declining new cases and rising cumulative cases (Fig. 3D), during unlock-4, advancing or delaying change-point onset by five days showed insignificant difference in cumulative cases (Fig. 3E). The cumulative cases remained unchanged with the change in transient duration of intervention, the new cases showed similar variation as *λ* (Fig. 3C and 3F).

**Fig. 3.**
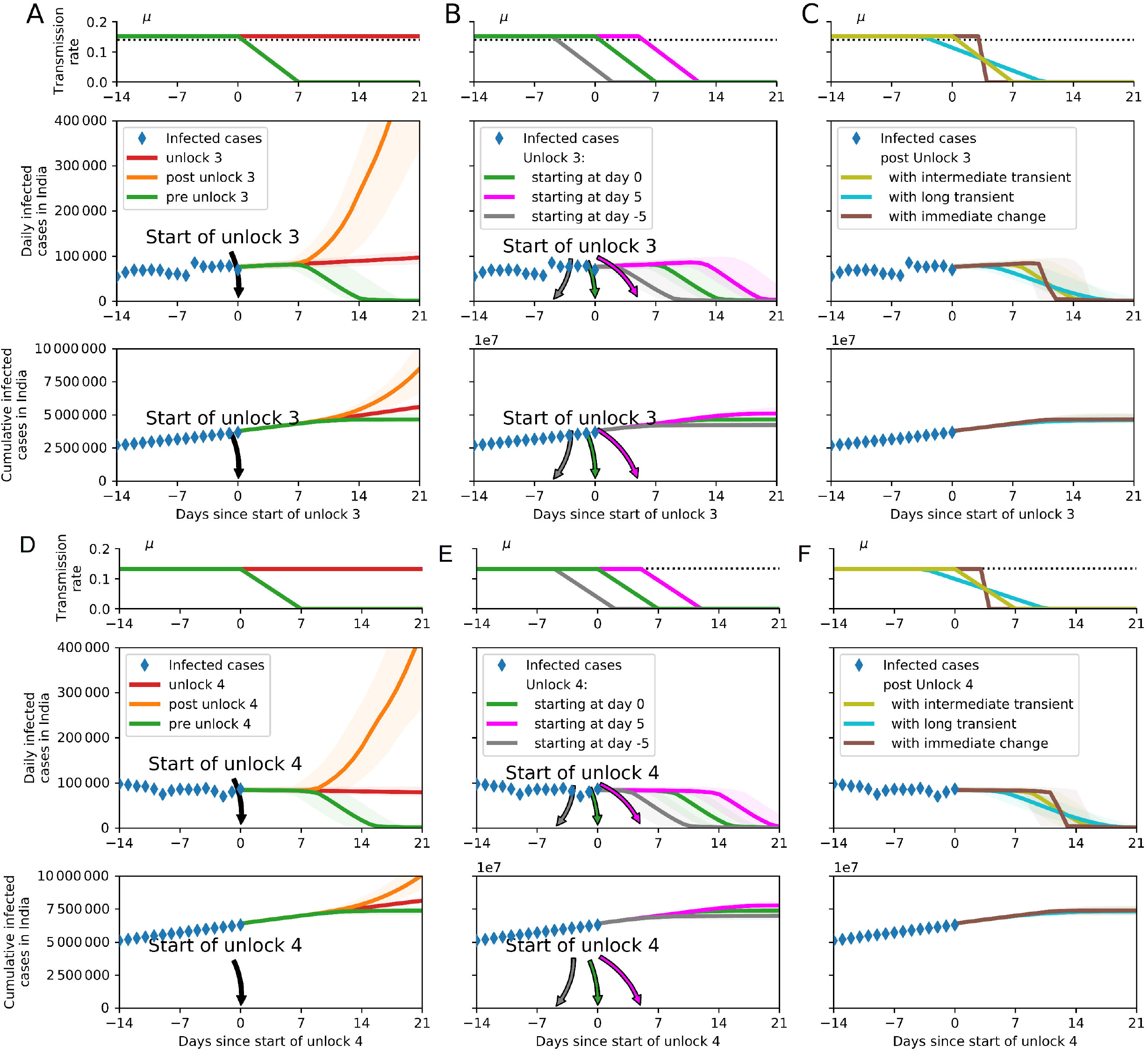
Effect of strength, timing and duration of unlock-3 and unlock-4 measures on infected case numbers. Each unlock measure featured re-opening of activities outside Containment Zones in phased manner and strict lockdown in containment zones only. Specifically unlock-3 removed night curfews, reopened recreational centres like gymnasiums and yoga centres. Unlock4 reopened metro rail in graded manner, and permitted limited gatherings. Under the extended relaxations, social distancing were hypothesized to be ∼ 0.9 factor stronger and ∼0.9 factor milder respectively, as a pre- and post-effect of unlock measure. **(A)** With respect to the strength of unlock-3, the transmission rate remained nearly the same but the daily and cumulative infected cases increased. Perpetuation of pre-unlock scenario would have caused decline in all. **(B)** Delaying the onset timings of unlock-3 measure showed insignificant change in cumulative case numbers: unlock-3 starting on 1 August 2020 (green), 5 days later (magenta), or 5 days earlier (gray). **(C)** The cumulative cases remained unchanged with change in transient [immediate (brown), intermediate (green), long (cyan)] duration of unlock-3, the new cases showed similar variation as λ. **(D), (E), (F)** same effect as **(A), (B), (C)** but with declining daily infected cases in unlock-4 starting 1 September 2020.

The SIR-model parameters with two identified change-points without weekend effect showed that the first change-point matched with the timelines of publicly announced strategies around 1 August 2020, when unlock-3 began, coinciding with continued closure of educational institutions and banned social gatherings, permission of interstate transport, besides release of night curfews. This change-point featured *λ*_1_ = 0.17 (0.12 − 0.21) that unfolded over 2.9 days (1.6 − 5.4) (Fig. 4F). The second change-point was detected around 1 September 2020, which coincided with the announcement of unlock-4, featured by lockdown measures remaining in force in containment zones, some activities permitted outside containment zones with reopening of metro-rail in graded manner, small gatherings permitted, continued compulsion of face-masking in public. The second change point had *λ*_2_ = 0.15 (0.10 − 0.19) that unfolded over 3.3 days (1.7 − 6.7) (Fig. 4F). With *λ*_0_ = 0.16, *σ* = 15.6, *D* = 9.6 days, *μ* = 0.15, the change-points were quantified as 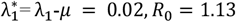, during unlock- and 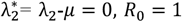, during unlock-4, implying effectiveness of unlock-4 measure bringing 100% reduction of *λ*^*^ and decline of the COVID-19 epidemic (Fig. 4). Thus the effectiveness of an intervention modelled as Bayesian change points could help us interpret the impact of different control measures and to include them into forecasts. Previously, Bayesian inference of COVID-19 change points correlating with social distancing restrictions were applied using the example of Germany (Dehning et al., 2020). Similar model was used to detect and assess latent events associated with spreading rates in South Africa (Mbuvha et al., 2020) and the US (Jiang et al., 2020).

**Fig. 4.**
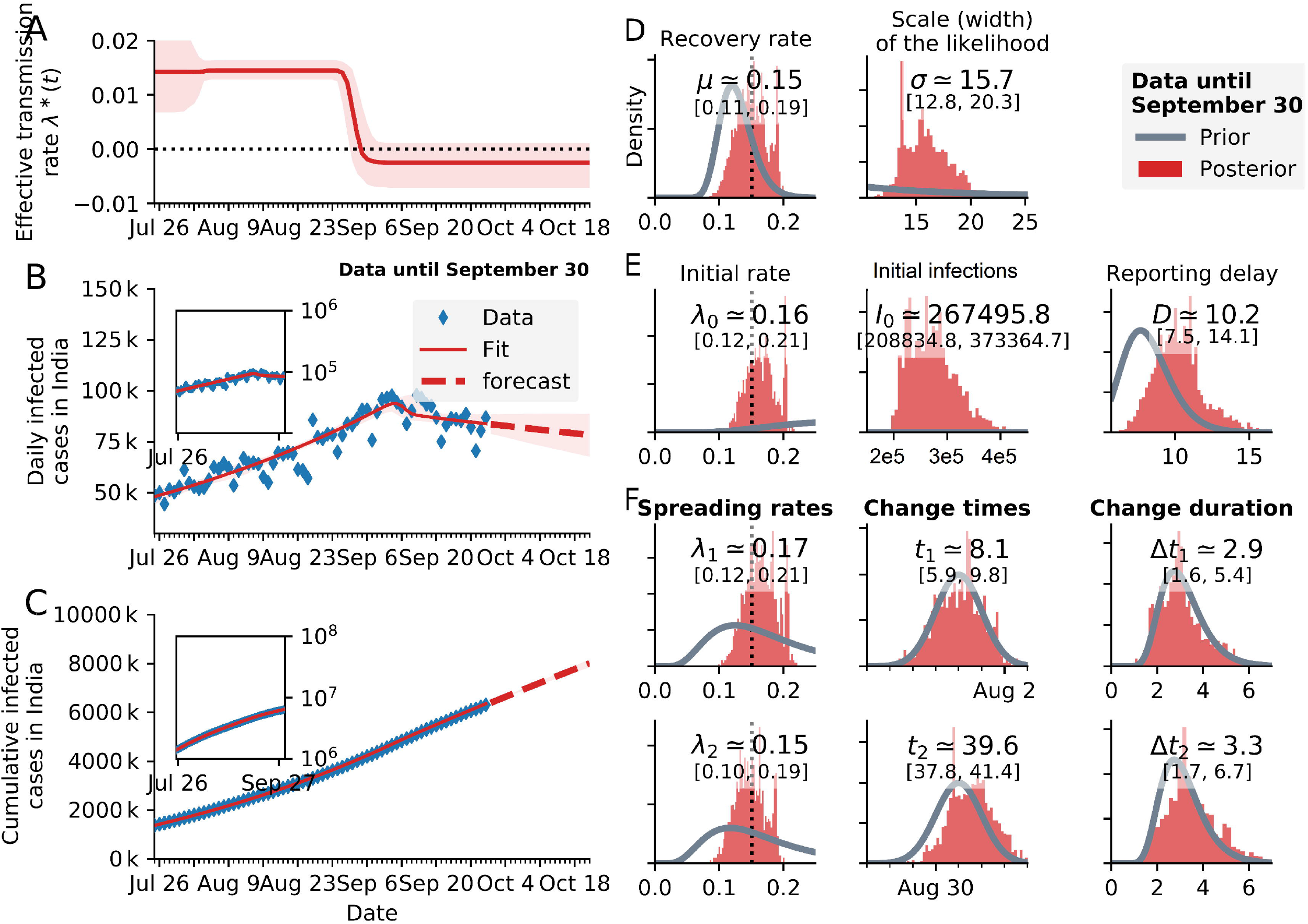
Identification and quantification of change points in COVID-19 transmission rate of India during unlock-3 and unlock-4 phase using SIR model without weekend effect. Time-series SIR model fit estimates of the **(A)** effective transmission rate *λ*^*\**^(*t*), **(B)** daily infected cases compared to the observed data, and **(C)** cumulative infected cases compared to the observed data. Inset shows semi-log plots. Underreporting factor on 18 October 2020, for daily and cumulative infected cases were and respectively, using SIR model. **(D-G)** Priors and posterior distribution of model parameters, values are expressed in median and 95% *CIs* of the posteriors. The SIR exhibited 5.97 lower LOO score than SEIR, both models with 2 change points and a weekend factor. Thus SIR model represented better consistency with observed data compared to SEIR model.

Compared to the two-change-point model without weekend-correction (*ELPD*-*LOO* = −687.67, *SE* = 8.00, *pLOO* = 6.71, computed from 2000 by 68 log-likelihood matrix) (Table 3), the *ELPD*-*LOO* for the SIR-model, also computed from 2000 by 68 log-likelihood matrix, with two change-points and weekend-modulation was higher by 30.76 (*ELPD*-*LOO* = −656.91, *SE* = 6.82, *pLOO* = 7.43) (Table 3), [*λ*_0_ = 0.16, *λ*_1_ = 0.17, *λ*_2_ = 0.15, *μ* = 0.15, *σ* = 10.2, *D* = 12.8days, 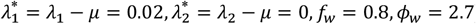], (Fig. 5). Sampling were run for chains with 1000 tunes and 500 draw iterations so that a total of 6000 draws occurred. Thus, in our study the lower *LOO* in the former, differing by 1.18*SE*, indicated higher consistency of the two-change-points SIR model with data excluding the weekend-factor that implied homogenous reporting of daily new cases through the entire week irrespective of weekend effect. However, higher number of COVID-19 daily new cases were reported during weekdays compared to weekends in Germany, substantiated by lower *LOO* score in weekend-effect model compared to that without weekend-effect model (Dehning et al., 2020).

**Fig. 5.**
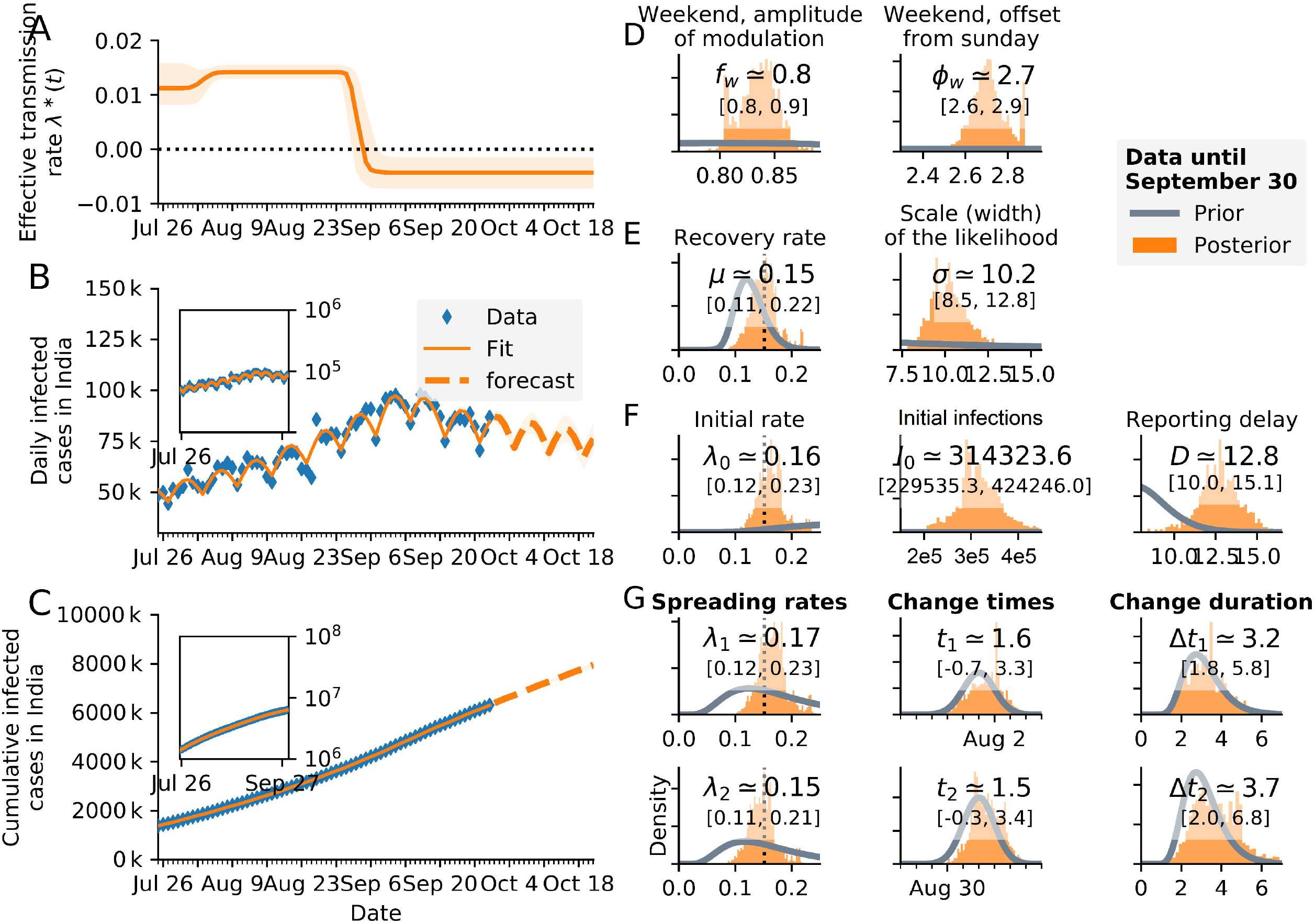
Identification and quantification of change points in COVID-19 transmission rate of India during unlock-3 and unlock-4 phase using SEIR model with weekend effect. Time-series SIR model fit estimates of the **(A)** *λ*^*^(*t*), **(B)** daily infected cases compared to the observed data, and **(C)** cumulative infected cases compared to the observed data. Inset shows semi-log plots. Underreporting factor on 18 October 2020, for daily infected cases were higher using SEIR model compared to SIR model whereas the underreporting factor for cumulative infected cases were same as SIR model. **(D-G)** Priors and posterior distribution of model parameters, values are expressed in median and 95% CIs of the posteriors. The SEIR model featured an additional incubation period *D*_*inc*_ with prior lognormal (5, 1) scale parameter 0.418, and an initial exposed function *E*_0 ∼_HalfCauchy(10). The corresponding priors for reporting delay *D* were (5, 0.2), *λ*_0_ (2, 0.7), *μ* (0.3, 0.3), and the other priors were same as SIR model.

Application of SEIR model based on Eqs. 4 and 5 with two-change-points and weekend-modulation as per Eqs. 6 and 7 (*f*_*w*_ = 0.8, *ϕ*_*w*_ = 2.7), exhibited greater negativity of the effective transmission rate with unlock-4 compared to unlock-3 measure 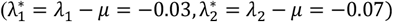 (Fig. 6), and higher transmission and recovery rates (*λ*_0_ = 0.36, *λ*_1_ = 0.30, *λ*_2_ = 0.26, *μ* = 0.33) (Fig. 6) compared to the SIR model (Fig. 5). Cross-validation showed the *ELPD*-*LOO* for the SEIR-model (−650.94, *SE* = 7.16, *pLOO* = 13.66, computed from 2000 by 68 log-likelihood matrix) slightly greater (by 5.97) than that of the corresponding SIR-model (Fig. 5), with 1*SE*(0.34) higher variation (Table 3), indicating considerably greater evidence for SIR model with respect to SEIR in explaining current COVID-19 data in Indian context. Similarly, SIR model displayed superior goodness of fit to the SEIR on South African data whereas SEIR produced a slightly better *LOO* score than the SIR main model on German data (Dehning et al., 2020; Mbuvha et al., 2020).

**Fig. 6.**
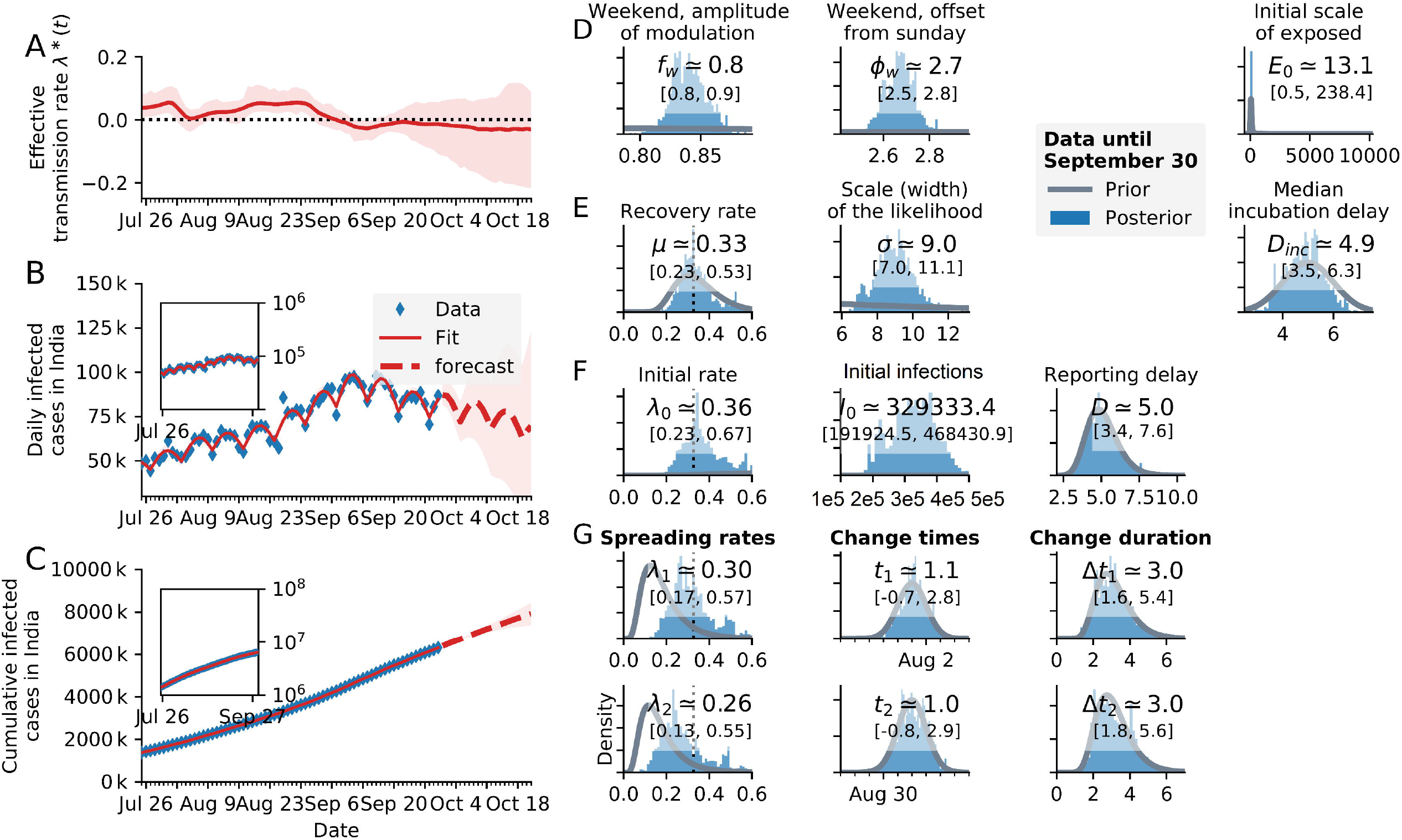
SIR model with two change points and weekend effect. Same as Fig. 4 but with weekend effect and higher LOO score.

Estimating the effect of change-points is vital for priors settings that help to anticipate the effects of any impending change points and accordingly make future projections. The SIR-model with one change point (Fig. 7) centred around the implementation of unlock-3 on 1 August 2020 showed superior goodness of fit with the COVID-19 observed data in India compared to the model with two change-points announcement of unlock-3 and unlock-4 on 1 August 2020 and 1 September 2020 respectively (Fig. 5); both models examined over the period from 25 July 2020 to 30 September 2020, with weekend-modulation. This was evident from the lower (by 43.14) *ELPD*-*LOO* score with one change point (−700.05, *SE* = 6.14, *pLOO* = 6.24) (Table 3), (*λ*_0_ = 0.17, and *λ*_1_ = 0.16, *μ* = 0.15, *σ* = 17.6, *f*_*w*_ = 0.8, *ϕ*_*w*_ = 2.6, *D* = 8.7 days, 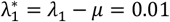 (Fig. 7) that fitted the observed data better compared to that with two change-points (Fig. 5) with < 1 *SE* (= 0.68) lower difference (Table 3). This was also clear from the simulation effect of hypothetical inventions on future COVID-19 cases in India, which showed that continuation of pre-unlock situation would have caused further decrease in both daily new and cumulative infected cases (Fig. 3A and 3D), implying that extension of stricter social-distancing measures would have been advantageous in reducing cases. Association of COVID-19 transmission in India in the context of containment measures demonstrated lower *ELPD*-*LOO* = −305.39 (*SE* = 4.58, *pLOO* = 3.45) for unlock-3, computed from 2000 by 30 log-likelihood matrix, and for the unlock-4 computed from 2000 by 29 log-likelihood matrix as -299.04 (*SE* = 4.08, *pLOO* = 2.75) (Table 3), using Eqs. 16 to 18. SIR models with three change-points described the data better than fewer change points on German data and SIR model with two change points was the best fit on South African data, as exhibited by the *LOO* cross-validation and all change points coinciding with respective government interventions (Dehning et al., 2020; Mbuvha et al., 2020). Surveillance of COVID-19 pandemic involved a reporting delay factor (range: 7 − 13 days) that was composed of testing delay between the incubation period of the virus (time period for the symptoms to develop following infection with the virus, with median estimates of 5 − 6 days) and the testing date (1 − 3 days); an additional delay occured between the testing date and results date (1 − 4 days) (Lauer et al., 2020). The D extrapolated from SIR-model with two change-points (12.8 days) (Fig. 5F) versus one change point (8.7 days) with weekend modulation (Fig. 7F), SIR model with two change points without weekend modulation (10.2days) (Fig. 5F), SEIR model with two change points with weekend modulation (reporting plus incubation delay 10.9 days) (Fig. 6F), indicate consistency with the above mentioned summated delay factor; the inherent difference within the obtained values might be due to the experimental conditions and model types adopted. The median change duration in all such situations were estimated to be around 3 days that were necessary to enact interventions, in the form of continuing and lifting of restrictions based on containment areas. Thus, the reporting delay combined with the change duration ranged from 11 to 16 days, which represented the time gap required to identify any change points in infected case numbers that in conjunction with the effective COVID-19 transmission rate, help to determine pertinent containment measures. The India COVID-19 daily infected case numbers using SIR and SEIR models were estimated to be around 75,000 and 72,000 respectively; the cumulative infected case numbers using both models were estimated at 8000,000 as of October 18, 2020 (for the period from 25 July 2020 to 30 September 2020); the effective transmission rate stabilized at less than zero, *R*_0_ = 1 and 0.79 using SIR and SEIR models respectively (Figs. 5 and 6).

**Fig. 7.**
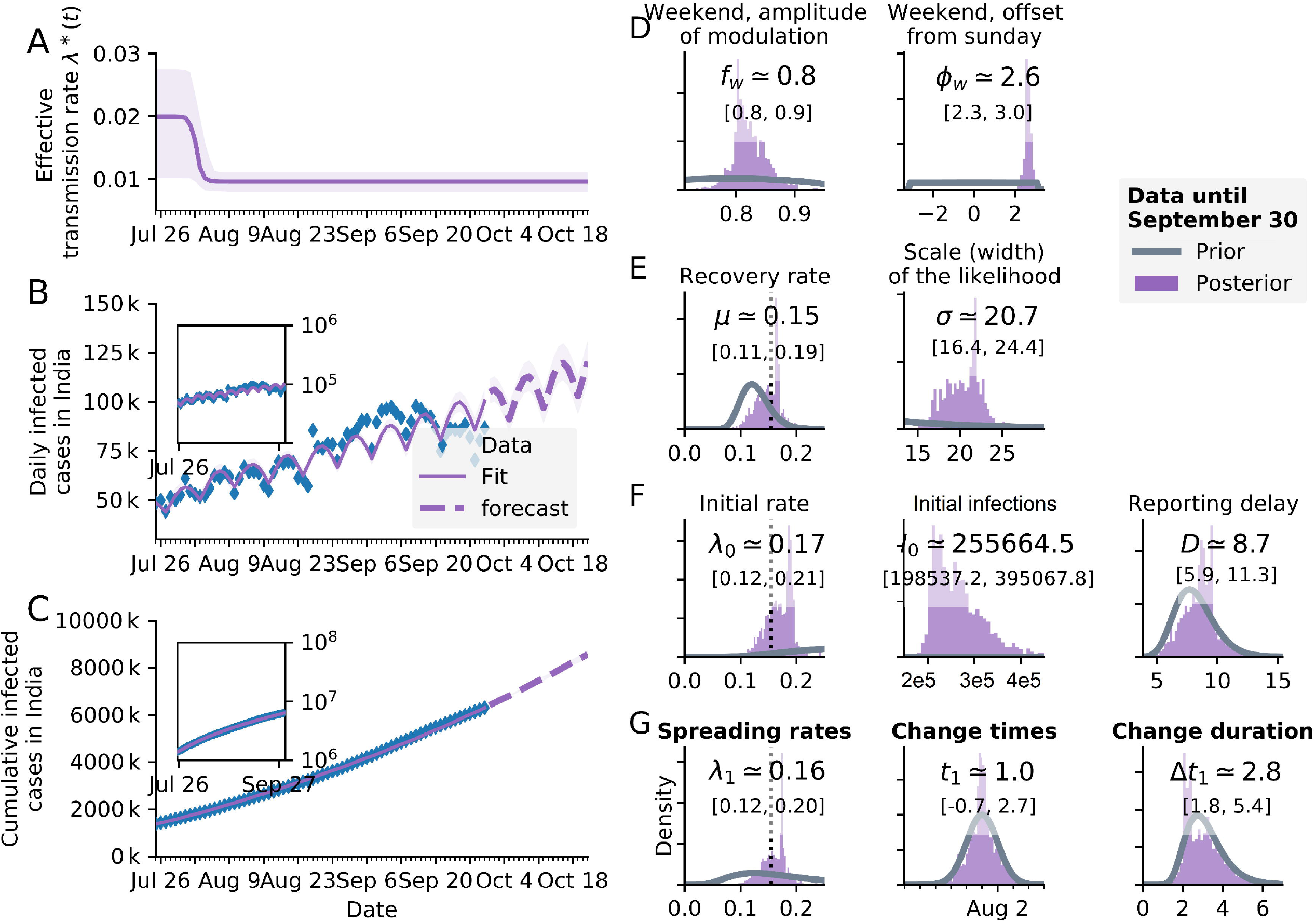
SIR model with one change point with weekend effect. Same as Fig. 4 but with one change point and weekend effect and lower LOO score. Most favoured model. Underreporting factor on 18 October 2020, for daily infected cases were higher 2.1 (=130,000/61,871) with this model compared to SIR model in Fig. 4.

## 4. Conclusion

Overall, the SIR model, including weekend-modulation and one-change point, with continuation of intervention similar to the unlock 3 situation was favoured over other models. However, the finding from the SIR model including weekend-modulation and two-change points that, *μ* was 0.15, *λ*^*^ was 10% and ‘zero’ during unlock-3 and unlock-4 respectively, implied unlock-4 measure brought 100% reduction of *λ*^*^, beginning around 5 September, 2020 (Fig. 5A-G), indicating new recoveries exceeding the new infections. Therefore, the epidemic curve is expected to decline to the baseline level, when the effective transmission rate becomes remarkably negative leading to sustained dwindling of new infections, provided no re-infection occurs and non-pharmaceutical interventions such as voluntary face-masking, physical-distancing, in addition to government measures including graded lockdown intervention in containment zones are maintained.

## Data Availability

Data used in this study are publicly available, and are preserved with the authors in this connection.

## Author’s contribution

Manisha Mandal and Shyamapada Mandal jointly designed the study, analysed and interpreted the data, discussed and wrote the manuscript.

